# ECG-derived global longitudinal strain using artificial intelligence: A comparative study with transthoracic echocardiography

**DOI:** 10.1101/2024.04.29.24306468

**Authors:** Hong-Mi Choi, Joonghee Kim, Jiesuck Park, Jun-Bean Park, Hyung-Kwan Kim, Hye Jung Choi, Yeonyee E. Yoon, Goo-Yeong Cho, Youngjin Cho, In-Chang Hwang

**Author notes:** **Address for correspondence:** Youngjin Cho, Cardiovascular Center, Seoul National University Bundang Hospital, 82 Gumi-ro-173-gil, Bundang, Seongnam, Gyeonggi 13620, South Korea, AND, In-Chang Hwang, Cardiovascular Center, Seoul National University Bundang Hospital, 82 Gumi-ro-173-gil, Bundang, Seongnam, Gyeonggi 13620, South Korea. **Tweet:** AI-based estimation of the GLS from ECG is a practical alternative to the LVGLS on echocardiography #Artificial intelligence; #Electrocardiography; #Global longitudinal strain.

## Abstract

**Background:** Despite the versatility of the left ventricular (LV) global longitudinal strain (LVGLS), its complex measurement and interpretation make it difficult to use. An artificial intelligence (AI)-generated electrocardiography (ECG) score for LVGLS estimation (ECG-GLS score) may offer a cost-effective alternative.

**Objectives:** We evaluated the potential of an AI-generated ECG-GLS score to diagnose LV systolic dysfunction and predict the prognosis of patients with heart failure (HF).

**Methods:** A convolutional neural network-based deep-learning algorithm was trained to estimate the echocardiography-derived GLS (LVGLS) using retrospective ECG data from a tertiary hospital (n=2,882). ECG-GLS score performance was evaluated using data from an acute HF registry at another tertiary hospital (n=1,186).

**Results:** In the validation cohort, the ECG-GLS score could identify patients with impaired LVGLS (≤12%) (area under the receiver-operating characteristic curve [AUROC], 0.82; sensitivity, 85%; specificity, 59%). ECG-GLS performance in identifying patients with an LV ejection fraction (LVEF) of <40% (AUROC, 0.85) was comparable to that for LVGLS (AUROC, 0.83) (p=0.08). Five-year outcomes (all-cause death; composite of all-cause death and hospitalization for HF) occurred significantly more frequently in patients with low ECG-GLS scores. Low ECG-GLS score was a significant risk factor for these outcomes after adjustment for other clinical risk factors and LVEF. The prognostic performance of the ECG-GLS score was comparable to that of the LVGLS.

**Conclusions:** The ECG-GLS score demonstrates a strong correlation with the LVGLS and is effective in risk stratification for the long-term prognosis after acute HF, suggesting its potential role as a practical alternative to the LVGLS.

**Condensed abstract:** This study is the first to attempt to estimate the left ventricular global longitudinal strain (LVGLS) from electrocardiography (ECG) data using an artificial intelligence-based algorithm (ECG-GLS score). The ECG-GLS score was correlated with the LVGLS and performed as well as the LVGLS in predicting the long-term prognosis of patients with heart failure. Thus, the ECG-GLS score has potential as practical alternative to the LVGLS on echocardiography, with reductions in time and effort.

**Graphical Abstract:** 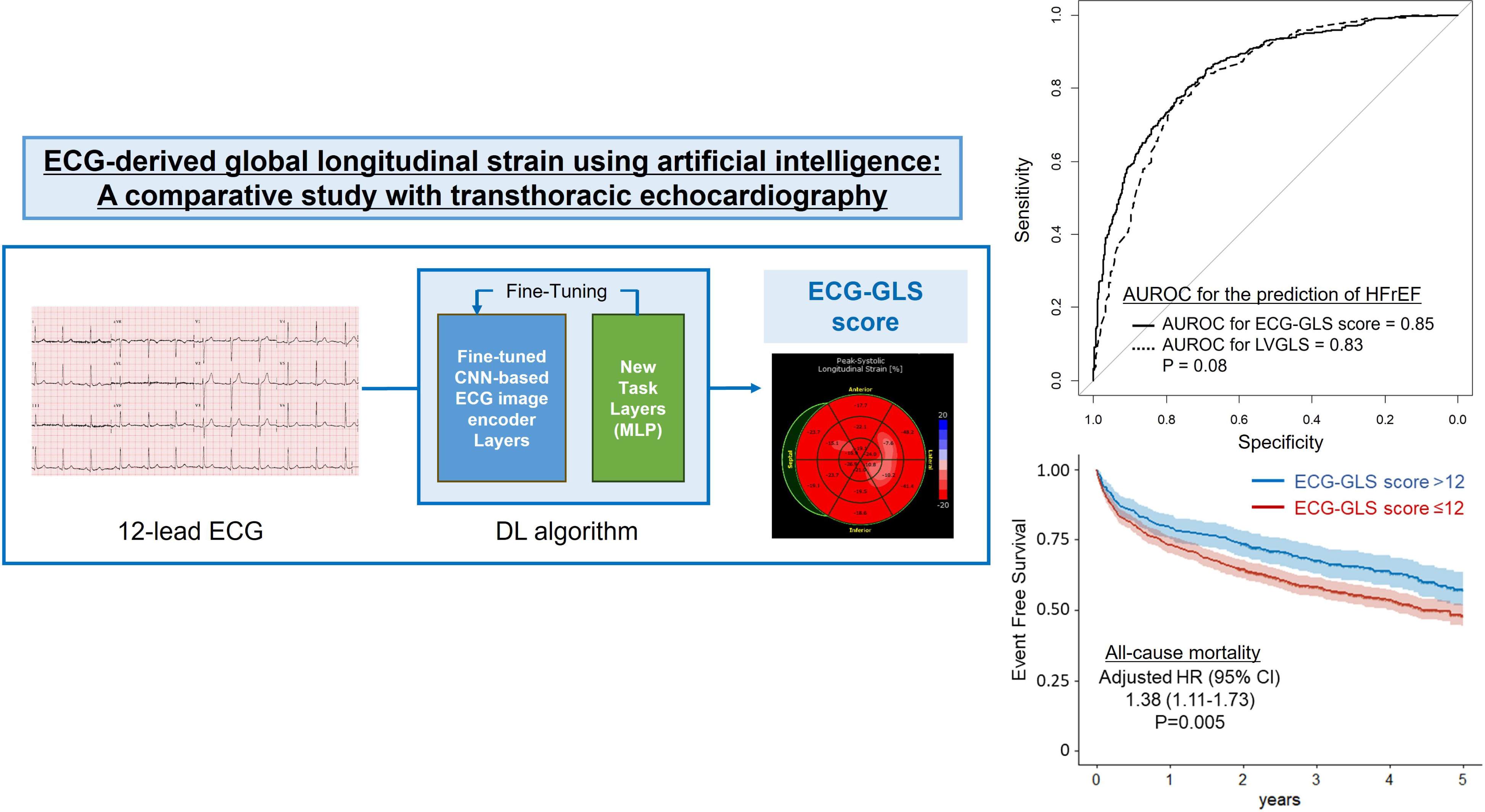

## INTRODUCTION

Recent developments in echocardiography have emphasized the significance of the left ventricular (LV) global longitudinal strain (GLS; LVGLS) as a prognosticator of various heart diseases,^1^ including heart failure (HF),^2^ valvular heart diseases,^3–5^ and cardiomyopathies.^6^ Despite its vast clinical utility, there are barriers to the widespread use of LVGLS. For instance, the measurement and interpretation of the LVGLS are time-consuming and complex compared to the ejection fraction; specific machines, software, and skilled personnel are required.

Electrocardiography (ECG) is a simple, cost-effective, and well-established method for heart-disease screening. Advances in artificial intelligence (AI) have expanded the capabilities of ECG, enhancing its diagnostic scope in the cardiovascular field.^7^ AI can also assist in the diagnosis of subclinical atrial fibrillation, HF, hypertrophic cardiomyopathy (HCM), and valvular heart diseases.^8^ Previous studies indicate potential in AI-assisted analysis of ECG patterns and features for not only the detection of certain diseases, but also for the estimation of LV function. For example, AI-enabled ECG analysis can distinguish ST-segment elevation myocardial infarction or LV systolic dysfunction among patients who visit the emergency department.^9,10^

Given the expanding clinical utility of LVGLS, with its superior sensitivity over that of the LV ejection fraction (LVEF) and robust prognostic value across various heart diseases, leveraging AI technology to estimate the LVGLS from ECG features has merit, as this could enhance its clinical efficacy and cost-effectiveness. In the present study, we generated an ECG-derived GLS (ECG-GLS) score using an AI deep-learning algorithm and evaluated its potential in diagnosing LV systolic dysfunction and predicting the prognosis of patients with HF.

## METHODS

### Study population and data management

This multicenter retrospective cohort study involved two tertiary hospitals (Hospitals A and B). ECG and echocardiographic data were obtained from the following four registries: 1) the STRATS-AHF cohort, which included patients who were admitted for acute HF with a range of ejection fractions from June 2009 to June 2015 (n=1608);^2^ 2) ARNI baseline cohort, which included patients with HF with reduced ejection fraction (HFrEF) who were prescribed ARNI from September 2015 to April 2020 (n=409); 3) ARNI follow-up cohort from December 2020 to January 2021 (n=409);^11^ and 4) three-chamber strain cohort, which included patients who underwent echocardiography with GLS data from February 2020 to November 2022, irrespective of diagnosis (n=984). The validation cohort comprised patients in the STRATS-AHF registry from Hospital B.^2^ A total of 2,882 and 1,186 echocardiography and ECG pairs were included in the training and validation cohorts, respectively (**Figure 1**). Echocardiography and 12-lead ECG pairs performed within a 7-day interval were included in the analysis.

**Figure 1.**
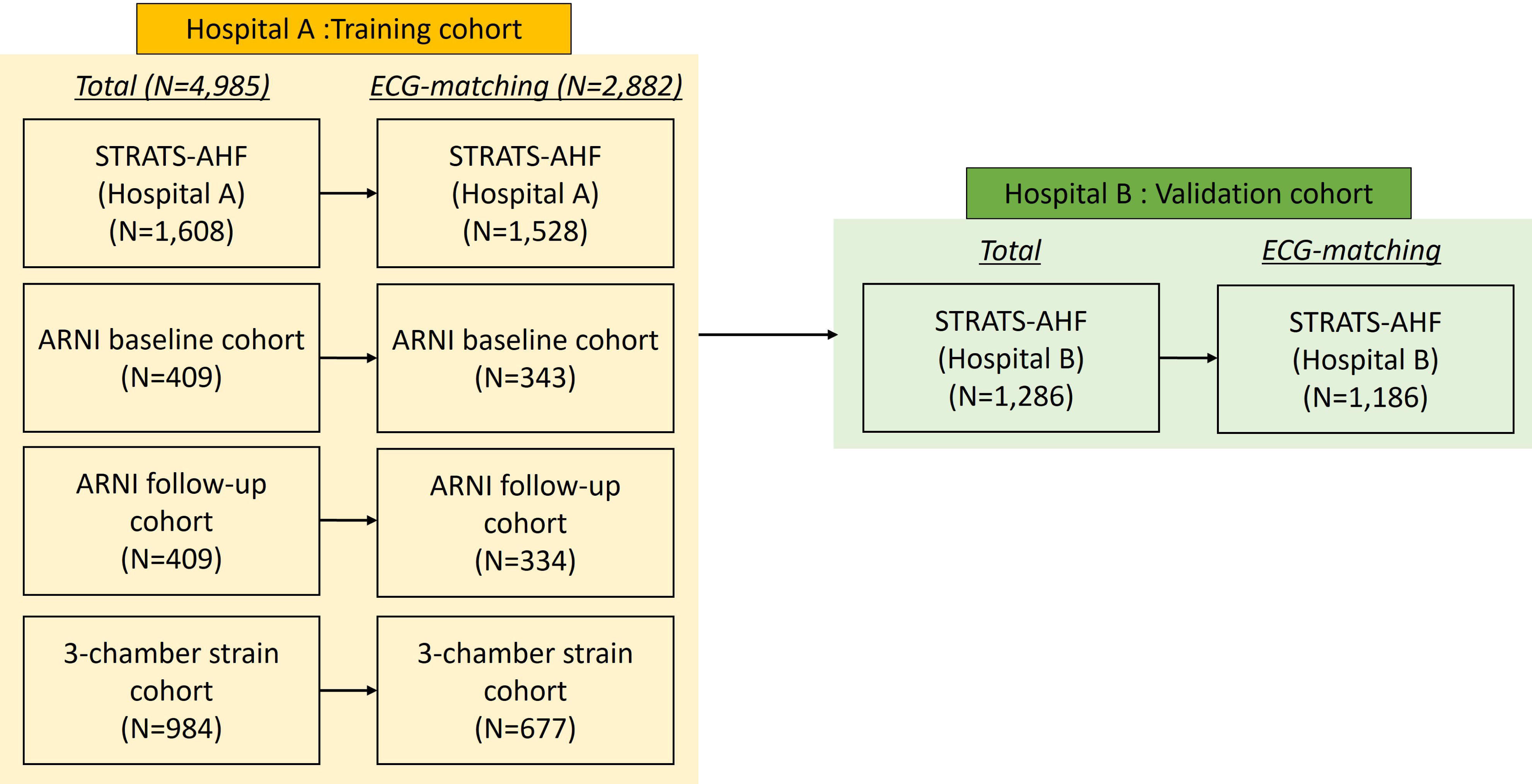
Composition of the training and validation cohorts. Echocardiography and 12-lead electrocardiography (ECG) pairs performed within a 7-day interval were included in the analysis.

Clinical data were collected by reviewing electronic medical records. The study outcomes were the 5-year all-cause death (ACD) and a composite of the 5-year ACD and hospitalization for HF (HHF).^2^

This study was approved by the Institutional Review Board of each hospital (hospital A, B-2212-801-102; hospital B, J-2302-117-1407). The requirement for informed consent was waived due to the retrospective nature of the study.

### AI algorithm

The AI algorithm was developed using a transfer-learning approach that leveraged the encoder component from an existing deep-learning system. This system, designed to classify cardiac rhythms and provide various risk scores for emergency conditions, utilizes a common ECG encoder based on a modified ResNet architecture and has received approval from the Korean Food and Drug Administration as a medical device.^9,12^ We extracted the encoder portion from the system and integrated it with two fully connected layers to generate a single numerical output for predicting the GLS. This integrated model was fine-tuned using an ADAM optimizer to minimize the mean squared error between the predicted and measured GLS values in the training dataset.

### Echocardiography and strain analysis

Echocardiographic images were obtained using comprehensive echocardiography in accordance with the guidelines of the American Society of Echocardiography.^13^ LVEF was measured from apical four- and two-chamber views using the Modified Simpson’s method. LVGLS was measured using a built-in software of each echocardiography machines. For the training cohort 2 and 3, Image-Arena system (TomTec Imaging Systems, Munich, Germany) was utilized to measure LVGLS. For strain analysis, the endocardial borders were traced on the end-systolic frame defined by the QRS complex. The software tracked speckles along the endocardial border and myocardium throughout the cardiac cycle. The peak longitudinal strain was automatically computed by averaging regional strain values. The LVGLS was obtained from apical three-, four-, and two-chamber views. Because the LVGLS values are expressed as absolute values to avoid confusion, higher values represent better function. For patients with sinus rhythm, analyses were performed on a single cardiac cycle. For patients with atrial fibrillation, strain values were calculated as the average of three cardiac cycles.

### Statistical analysis

Continuous variables are presented as means and standard deviations or medians with interquartile ranges and were compared using the independent two-sample t-test or Mann-Whitney U test. Categorical variables are presented as frequencies with percentages and were compared using the χ^2^ test. The primary measure of model performance was the area under the receiver operating characteristic curve (AUROC) for the prediction of LV systolic dysfunction estimated by the LVGLS. AUROC curves were compared using DeLong’s test.

Kaplan-Meier curves for study outcomes were plotted to indicate the discrimination capacities of the ECG-GLS score and LVGLS, which were compared using the log-rank test. We used Cox proportional hazards modeling to compare the predictability for study outcomes between the ECG-GLS score and the echocardiography-derived GLS (LVGLS). The hazard ratio (HR) was adjusted for parameters that showed an association with study outcomes on univariate analysis (p<0.1) or had relevant clinical significance, excluding those with >10% missing data or multicollinearity with other variables. The Harrell’s C-index (C-index) was calculated to compare the predictive performance of Cox proportional hazards models using the bootstrapping method. A two-sample t-test was conducted on 1,000 sets of bootstrapped C-indices to compare the two models.

Statistical significance was defined as a two-sided p-value <0.05. All statistical analyses were conducted using R software, version 4.1.2 (https://www.R-project.org).

## RESULTS

### Baseline characteristics and echocardiographic parameters

A total of 2,882 and 1,186 patients were included in the training and validation cohorts, respectively. The baseline characteristics are presented in **Table 1**. The training cohort comprised four registries containing patients with acute HF, chronic HF, various heart diseases, and normal heart function from Hospital A. The validation cohort was also drawn from one of the four registries, but the patients were from Hospital B. Age and sex were comparable between the training and validation cohorts; however, other features, including comorbidities, N-terminal pro-B-natriuretic peptide levels, and echocardiographic parameters, were significantly different. The mean interval between ECG and echocardiography was 0.58±1.26 and 1.00±2.49 days in training and validation cohorts, respectively.

**Table 1.** Baseline Characteristics of Training and Validation Cohorts.

|  | Training cohort 1 | Training cohort 2 | Training cohort 3 | Training cohort 4 |  | Training cohort (total) | Validation cohort |  |
| --- | --- | --- | --- | --- | --- | --- | --- | --- |
|  | STRATS-AHF (Hospital A) | ARNI cohort (initial) | ARNI cohort (follow-up) | All-comers cohort | P-value (Training Cohort 1-4) | Hospital A | Hospital B (STRATS-AHF) | P-value (Training vs. Validation) |
|  | (N=1528) | (N=343) | (N=334) | (N=677) |  | (N=2882) | (N=1186) |  |
| Age (years) | 72.3±13.1 | 64.6±13.7 | 64.8±14.1 | 68.4±14.4 | <0.001 | 69.6±13.9 | 69.0±14.8 | 0.246 |
| Male sex | 763 (49.9%) | 252 (73.5%) | 244 (73.1%) | 403 (59.5%) | <0.001 | 1662 (57.7%) | 681 (57.4%) | 0.912 |
| SBP (mmHg) | 133.4±29.1 | 119.1±18.0 | 118.4±17.4 | 128.2±18.8 | <0.001 | 128.8±25.3 | 129.0±27.4 | 0.826 |
| DBP (mmHg) | 74.8±18.6 | 71.7±13.9 | 71.2±3.1 | 73.7±12.7 | <0.001 | 73.8±16.3 | 75.9±16.6 | <0.001 |
| DM | 587 (38.4%) | 118 (34.4%) | 114 (34.1%) | 158 (23.3%) | <0.001 | 977 (33.9%) | 346 (29.2%) | 0.004 |
| HTN | 1049 (68.7%) | 110 (32.1%) | 108 (32.3%) | 271 (40.0%) | <0.001 | 1538 (53.4%) | 522 (44.0%) | <0.001 |
| Prior HF | 450 (29.5%) | 343 (100.0%) | 334 (100.0%) | 225 (33.2%) | <0.001 | 1352 (46.9%) | 812 (68.5%) | <0.001 |
| Prior CAD | 523 (34.2%) | 114 (33.2%) | 108 (32.3%) | 196 (29.0%) | 0.111 | 941 (32.7%) | 305 (25.7%) | <0.001 |
| BUN (mg/dL) | 27.6±18.0 | 22.3±12.7 | 22.2±12.6 | 20.6±13.3 | <0.001 | 24.7±16.1 | 26.1±17.0 | 0.023 |
| Creatinine (mg/dL) | 1.6±1.8 | 1.3±1.3 | 1.3±1.1 | 1.2±1.4 | <0.001 | 1.5±1.6 | 1.7±2.0 | <0.001 |
| LVEDD (mm) | 53.0±9.2 | 60.5±7.7 | 56.4±8.8 | 48.6±7.3 | <0.001 | 53.2±9.3 | 56.1±9.8 | <0.001 |
| LVESD (mm) | 40.3±11.3 | 51.1±8.7 | 45.3±10.3 | 33.4±8.9 | <0.001 | 40.5±11.6 | 41.7±12.3 | 0.012 |
| LVEF (%) | 41.5±15.8 | 27.7±6.8 | 38.0±11.9 | 56.7±12.2 | <0.001 | 43.0±16.3 | 40.7±15.4 | <0.001 |
| LVMI (g/m<sup>2</sup>) | 135.0±44.2 | 140.5±37.8 | 124.6±35.1 | 104.8±35.3 | <0.001 | 127.2±42.6 | 126.6±42.3 | 0.679 |
| LA diameter (mm) | 43.7±8.9 | 44.2±7.8 | 41.8±8.6 | 40.3±8.1 | <0.001 | 42.7±8.7 | 48.9±10.5 | <0.001 |
| LAVI (ml/m<sup>2</sup>) | 59.2±32.3 | 61.6±28.3 | 53.6±29.1 | 46.1±26.0 | <0.001 | 55.8±30.6 | 64.0±53.8 | <0.001 |
| E velocity (m/s) | 0.9±0.4 | 0.8±0.3 | 0.7±0.3 | 0.8±0.3 | <0.001 | 0.8±0.3 | 0.9±0.4 | <0.001 |
| E/e' | 19.1±10.8 | 18.9±10.5 | 15.0±9.0 | 13.0±7.3 | <0.001 | 17.1±10.1 | 19.4±11.0 | <0.001 |
| TR Vmax (m/s) | 2.8±0.6 | 2.7±0.6 | 2.5±0.5 | 2.5±0.5 | <0.001 | 2.7±0.6 | 3.0±0.6 | <0.001 |
| RVSP (mmHg) | 38.6±13.7 | 36.9±15.0 | 31.0±11.8 | 31.8±11.4 | <0.001 | 36.0±13.5 | 48.6±15.3 | <0.001 |
| LV GLS (% absolute) | 10.5±4.8 | 10.2±3.2 | 12.8±4.4 | 16.4±4.5 | <0.001 | 12.1±5.2 | 10.4±5.2 | <0.001 |
| Vendor |  |  |  |  | <0.001 |  |  | <0.001 |
| GE | 964 (63.3%) | 0 (0.0%) | 0 (0.0%) | 168 (24.8%) |  | 1132 (51.5%) | 547 (46.2%) |  |
| Philips | 167 (11.0%) | 343<br>(100.0%) | 334<br>(100.0%) | 509 (75.2%) |  | 676 (30.7%) | 266 (22.4%) |  |
| Siemens | 392 (25.7%) | 0 (0.0%) | 0 (0.0%) | 0 (0.0%) |  | 392 (17.8%) | 372 (31.4%) |  |

BUN = blood urea nitrogen; CAD = coronary artery disease; DBP = diastolic blood pressure; GLS = global longitudinal strain; HF = heart failure; LA = left atrium; LAVI = left atrial volume index; LVEDD = left ventricular end-diastolic dimension; LVEF = left ventricular ejection fraction; LVESD = left ventricular end-systolic dimension; LVMI = left ventricular mass index; NT-proBNP = N-terminal pro-B-type natriuretic peptide; SBP = systolic blood pressure; TR = tricuspid regurgitation.

### ECG-GLS performance

The LVGLS and ECG-GLS score were significantly correlated, with a correlation coefficient of 0.64 (p<0.001, **Figure 2A**). In the training cohort, the AUROCs of the ECG-GLS scores for detecting LVGLS ≤16% (indicating LV systolic dysfunction) and LVGLS ≤12% (indicating more severe LV systolic dysfunction) were 0.93 and 0.90, respectively. In the validation cohort, the AUROCs of the ECG-GLS score for detecting LVGLS ≤16% and LVGLS <12% were 0.85 and 0.82, respectively (**Figure 2B**). The precision, recall, and F1 score of the predictive model using the ECG-GLS score to identify patients with LVGLS ≤12% were 0.80, 0.84, and 0.82, respectively. The area under the precision-recall curve was 0.90 (**Supplementary Figure 1**). The diagnostic performance of the ECG-GLS score for identifying HFrEF (LVEF <40%) was comparable to that for the LVGLS (AUROC, 0.85 vs. 0.83, respectively; p=0.08; **Figure 3**).

**Figure 2.**
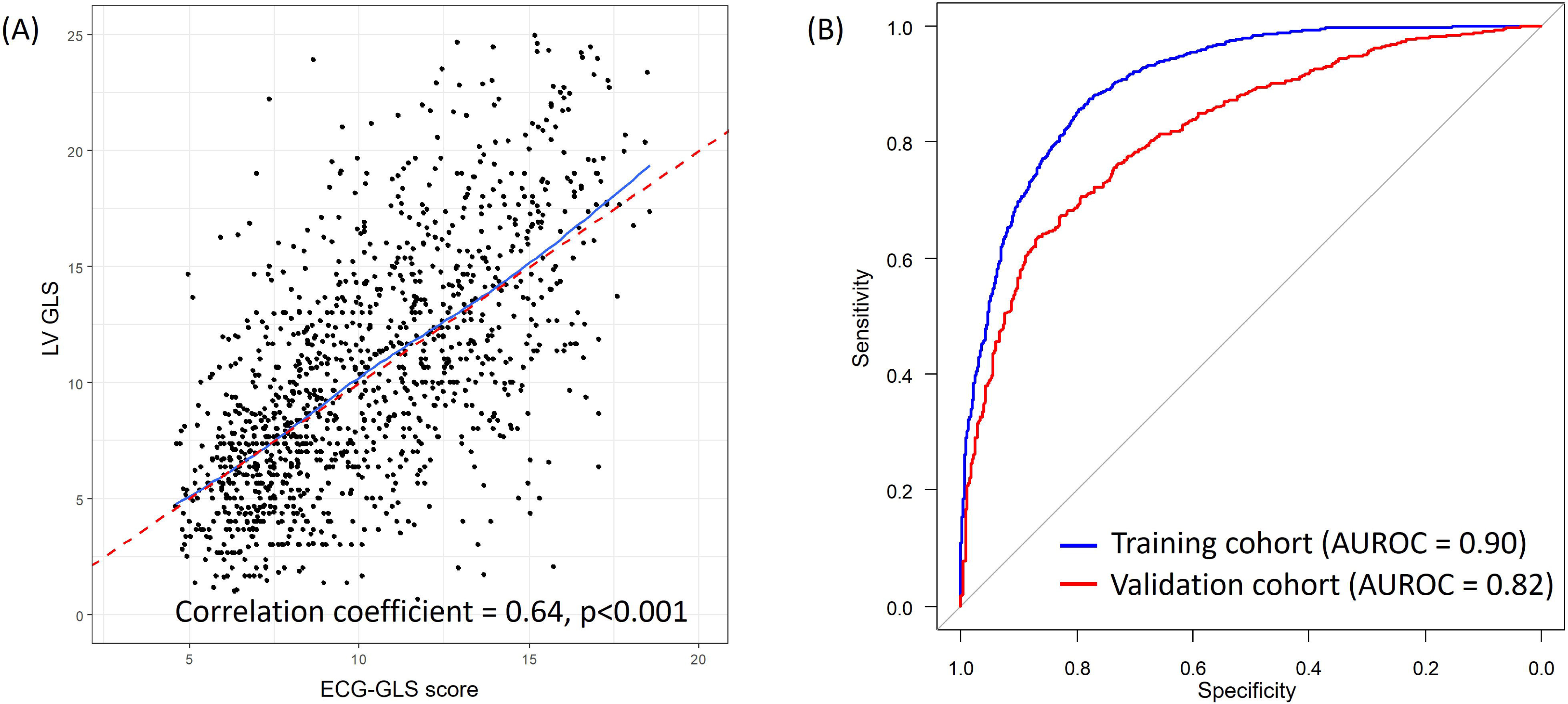
ECG-GLS evaluations. (A) The LVGLS and ECG-GLS score were significantly correlated (correlation coefficient=0.64, p<0.001). (B) Receiver-operating characteristic curves of the ECG-GLS score for the prediction of LVGLS ≤12% in the training cohort (blue) and validation cohort (red) are shown. Abbreviations: AUROC, area under the receiver operating characteristic curve; ECG-GLS, electrocardiography-derived global longitudinal strain; LVGLS, left ventricular global longitudinal strain on echocardiography

**Figure 3.**
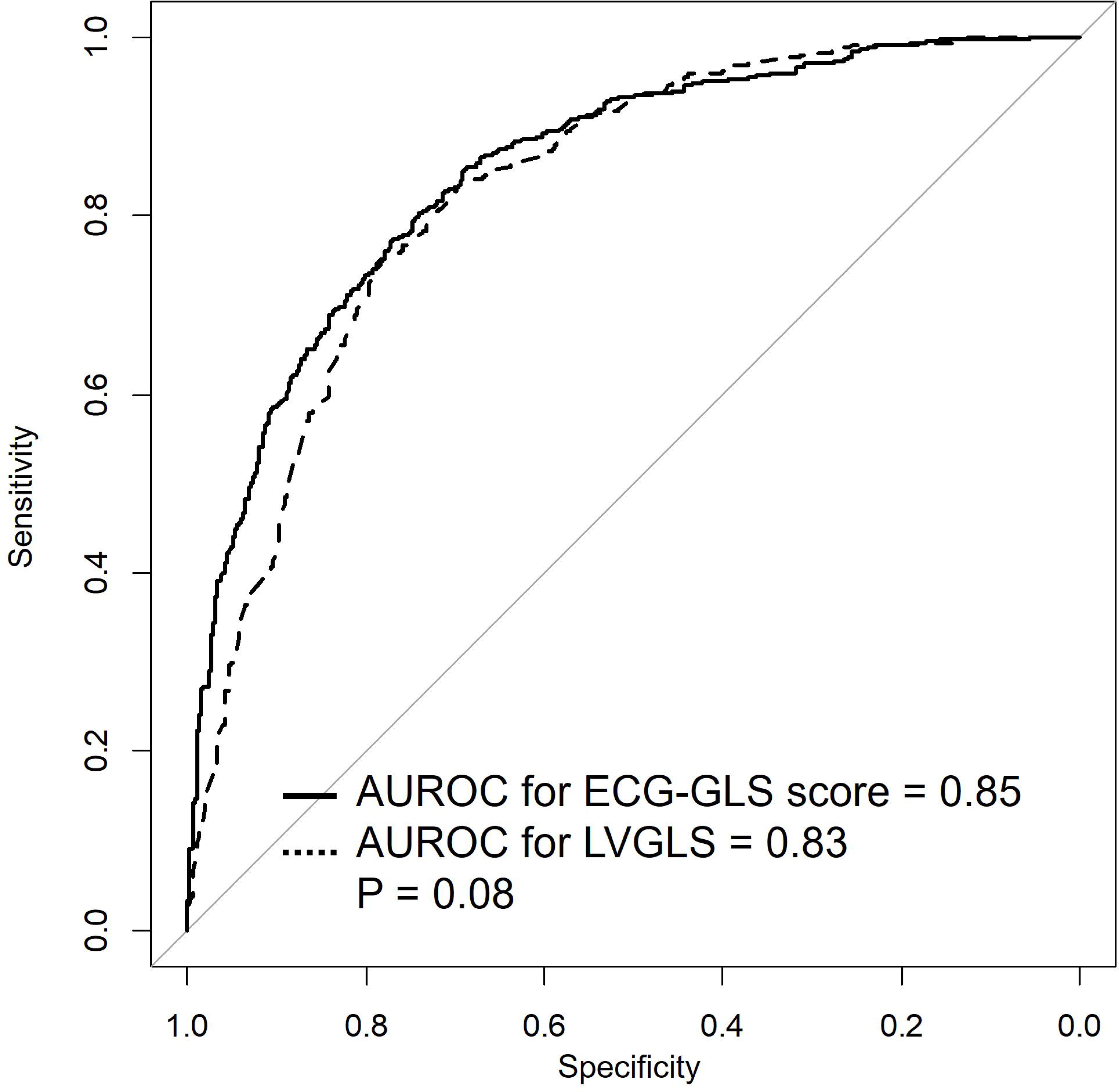
Receiver-operating characteristic curves. The performance of the ECG-GLS score (solid line) and LVGLS (dashed line) for the prediction of heart failure with reduced ejection fraction are shown. Abbreviations: AUROC, area under the receiver operating characteristic curve; ECG-GLS, electrocardiography-derived global longitudinal strain; LVGLS, left ventricular global longitudinal strain on echocardiography

### Prediction of study outcomes

Five-year ACD and the composite of ACD and HHF were significantly higher in patients with ECG-GLS scores ≤12 than in patients with ECG-GLS scores >12 (log-rank p=0.002 and p<0.001, respectively; **Figure 4A**). An analogous pattern was observed for the Kaplan-Meier curves using an LVGLS cutoff of 12% (log-rank p<0.001 and p=0.001, respectively; **Figure 4B**).

**Figure 4.**
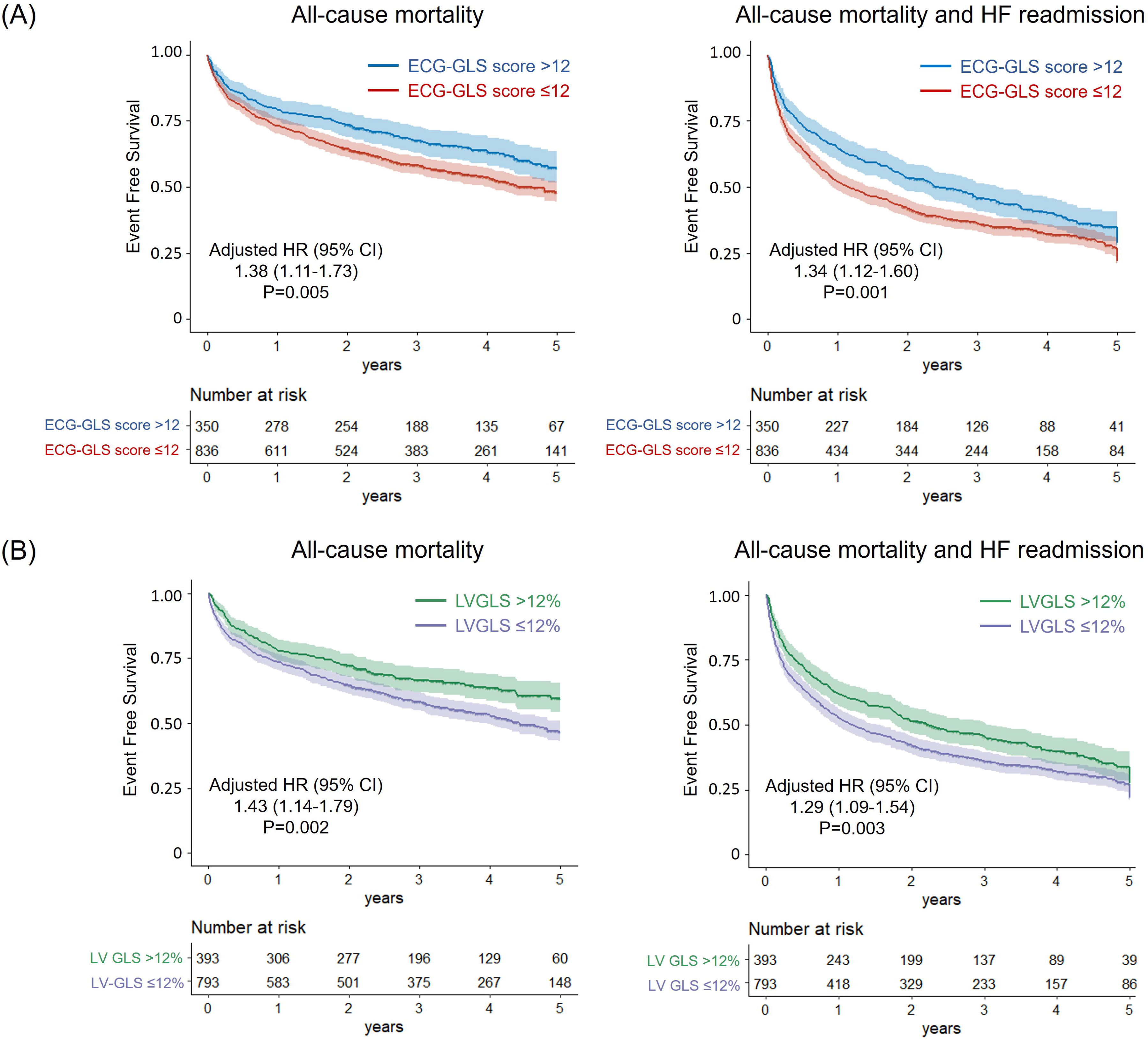
Survival curve comparisons. (A) Event-free survival curves for all-cause death and the composite outcome of all-cause death and HHF according to an ECG-GLS score >12 (blue) and ≤12 (red). (B) Event-free survival curves for all-cause death and the composite outcome of all-cause death and HHF according to an LVGLS >12% (green) and ≤12% (purple). Abbreviations: ECG-GLS, electrocardiography-derived global longitudinal strain; HHF, hospitalization for heart failure; LVGLS, left ventricular global longitudinal strain on echocardiography

LVGLS ≤12% (HR 1.41, 95% confidence interval [CI] 1.16-1.71) and ECG-GLS score ≤12 (HR 1.36, 95% CI 1.12-1.66) were associated with significantly higher 5-year ACD on univariable analysis (**Supplementary Table 1**). After adjustment for relevant clinical risk factors and echocardiographic parameters, low ECG-GLS score (≤12) was a significant predictor of ACD (HR 1.38, 95% CI 1.11-1.73), with comparable results to that for LVGLS ≤12% (HR 1.43, 95% CI 1.14-1.79; p-value for bootstrapped mean of Harrel’s C-indices=0.109; **Table 2**). Similar results were observed for the composite outcome of the 5-year ACD and HHF. LVGLS ≤12% (HR 1.29, 95% CI 1.09-1.54) and ECG-GLS score ≤12 (HR 1.34, 95% CI 1.12-1.60) were both significant indicators of the composite outcome (p-value for bootstrapped mean of Harrell’s C-indices=0.423; **Table 3**; univariable analysis results are provided in **Supplementary Table 2**).

**Table 2.** Multivariable Cox Proportional Hazard Regression Analysis for 5-Year All-Cause Death.

|  | Model 1 (LV GLS) |  |  | Model 2 (ECG-GLS) |  |  |
| --- | --- | --- | --- | --- | --- | --- |
|  | HR | 95% CI | P value | HR | 95% CI | P value |
| Age (years) | 1.04 | 1.03-1.05 | <0.001 | 1.04 | 1.03-1.05 | <0.001 |
| Male sex | 1.08 | 0.89-1.31 | 0.434 | 1.08 | 0.89-1.31 | 0.445 |
| DM | 1.32 | 1.09-1.59 | 0.004 | 1.34 | 1.11-1.61 | 0.003 |
| Creatinine (mg/dL) | 1.09 | 1.04-1.13 | <0.001 | 1.08 | 1.04-1.13 | <0.001 |
| Beta-blocker | 0.70 | 0.57-0.84 | <0.001 | 0.69 | 0.57-0.84 | <0.001 |
| RAS blocker | 0.71 | 0.59-0.85 | <0.001 | 0.69 | 0.57-0.83 | <0.001 |
| LVEF ≤30% | 1.21 | 0.97-1.50 | 0.090 | 1.24 | 1.00-1.53 | 0.052 |
| LVGLS ≤12% | 1.43 | 1.14-1.79 | 0.002 | | | |
| ECG-GLS score ≤12 | | | | 1.38 | 1.11-1.73 | 0.005 |

CI = confidence interval; HR = hazard ratio; RAS = renin-angiotensin system; other abbreviations as in Table 1.

**Table 3.** Multivariable Cox Proportional Hazard Regression Analysis for 5-Year All-Cause Death and Hospitalization for Heart Failure.

|  | Model 1 (LV GLS) |  |  | Model 2 (ECG-GLS) |  |  |
| --- | --- | --- | --- | --- | --- | --- |
|  | HR | 95% CI | P value | HR | 95% CI | P value |
| Age (years) | 1.03 | 1.03-1.04 | <0.001 | 1.03 | 1.03-1.04 | <0.001 |
| Male sex | 1.07 | 0.92-1.25 | 0.391 | 1.06 | 0.91-1.24 | 0.450 |
| DM | 1.39 | 1.19-1.61 | <0.001 | 1.39 | 1.19-1.61 | <0.001 |
| Creatinine (mg/dL) | 1.07 | 1.04-1.11 | <0.001 | 1.07 | 1.03-1.10 | <0.001 |
| Beta-blocker | 0.89 | 0.76-1.03 | 0.125 | 0.89 | 0.76-1.03 | 0.115 |
| RAS blocker | 0.84 | 0.72-0.98 | 0.023 | 0.82 | 0.71-0.96 | 0.011 |
| LVEF ≤30% | 1.08 | 0.90-1.28 | 0.409 | 1.08 | 0.91-1.29 | 0.360 |
| LVGLS ≤12% | 1.29 | 1.09-1.54 | 0.003 |  |  |  |
| ECG-GLS score ≤12 |  |  |  | 1.34 | 1.12-1.60 | 0.001 |

Abbreviations as in Table 1 and 2.

## DISCUSSION

In the present study, we demonstrated a strong correlation between the ECG-GLS (estimated from ECG features using an AI-based algorithm) and the LVGLS (measured using echocardiography). We also found that the performance of ECG-GLS for predicting the long-term prognosis of patients with HF and LVGLS was similar to that for the LVGLS, suggesting the relevance of our AI-based algorithm for estimating GLS from ECG features and its potential role as a practical alternative to the LVGLS. To date, the present study is the first to attempt to estimate the LVGLS from ECG data using an AI-based algorithm.

Research on AI-based algorithms that use ECG images to detect a range of heart diseases has been published, expanding the role of AI in cardiology.^8^ The utilization of AI-based ECG algorithms has focused on the early detection of diseases that can be easily diagnosed using ECG but are often missed, such as atrial fibrillation^14^ or ST-segment elevation myocardial infarction.^12^ However, the rapid growth of AI has expanded the potential of ECG to an extent that rivals that of echocardiography by revealing previous unseen pieces of information. Acceptable AI-enabled ECG performance has been demonstrated in the diagnosis of various types of HF^10,15,16^ and myocardial diseases, such as HCM^17^ and cardiac amyloidosis.^18^ Moreover, AI-enabled ECG HCM scores were correlated with decreases in LV outflow tract gradients and N-terminal pro-B-natriuretic peptide levels over time in patients with obstructive HCM who were prescribed mavacamten.^19^ Most recently, Lee et al. showed improved performance in the prediction of the prognosis according to diastolic dysfunction with AI-enabled ECG.^20^ In the present study, we attempted to extend the capability of AI-enabled ECG to predict the LVGLS, a state-of-the-art technology in the echocardiography field. Using ECG features of 2,881 patients from four retrospective registries, our AI-based algorithm for the estimation of LVGLS produced ECG-GLS scores that showed a good correlation with echocardiographic measurements of the LVGLS and were able to predict the long-term prognosis in patients who were admitted for HF.

In the present study, the AUROCs of the ECG-GLS score for detecting LVGLS ≤16% and ≤12% were 0.85 and 0.82, respectively, which were not as high as we expected. Therefore, the validation cohort comprised patients admitted for HF, whereas patients with all ranges of LVEF and LVGLS were included in the training cohort. In addition, the number of patients included in our study was smaller than that in other studies on AI-enabled ECG because of the limited collection of GLS data. Nonetheless, this study is the first to predict LVGLS using AI-enabled ECG. Our ECG-GLS score is comparable to the LVGLS in identifying patients with HFrEF. Furthermore, the ECG-GLS score is consistent with the familiar unit of the LVGLS. Thus, the ECG-GLS score is user-friendly and can be used interchangeably with the LVGLS in real-world settings.

Furthermore, the ECG-GLS score was comparable to the LVGLS score in predicting the long-term prognosis in patients. LVGLS has been shown to be a better indicator than LVEF for predicting cardiovascular outcomes in hospitalized patients with acute HF.^2^ Previous studies on AI-enabled ECG have predominantly emphasized its diagnostic capabilities, specifically for detecting various heart diseases or conditions. Although the diagnostic potential of AI-enabled ECG is considerable and holds significant promise for clinical use, augmenting the output of AI-enabled ECG algorithms with prognostic information can further enhance their utility. This integration of prognostic data reinforces and validates the diagnostic accuracy of the algorithm. Consistent with this, our findings indicate that AI-enabled ECG serves not only as an accurate indicator of the LVGLS, but also as a valuable tool for prognostication, thereby extending its role beyond mere diagnostics.

ECG has many advantages: it is ubiquitous, inexpensive, rapid, and does not require special training. Recently, AI researchers have expanded the role of ECG in the diagnosis of various heart diseases.^8^ Although the LVGLS has recently been recognized as a valuable tool, echocardiographic LVGLS measurements require additional time and effort from experts, as well as dedicated software or machines. The LVGLS also has the troublesome issue of between-vendor variability.^21^ Moreover, measuring the LVGLS using echocardiography may be limited in patients with poor echocardiographic windows, atrial fibrillation, and videos with low frame rates.^22^ In contrast, the ECG-GLS score is free from almost all of these limitations. Using the ECG-GLS score, non-experts can also predict the value of the LVGLS, diagnose LV systolic dysfunction, and forecast the outcome of patients easily, making it usable outside the echocardiography laboratory and significantly reducing time and costs. Integrating AI in interpreting ECG results could potentially improve the cardiac diagnostic process, making it more accessible and less reliant on specialized training and equipment.

### Limitations

This study has several limitations. First, the ECG and echocardiography pairs used in this study were insufficient to provide better prediction. However, the registries used in the training process have been shown to be useful in predicting the prognosis of patients with HF in previous studies. Moreover, the addition of more patients to the training cohort would require additional resources. Second, although various vendors were used in the measurement of the LVGLS, the number of ECG-echocardiography pairs used in the training cohort significantly differed among vendors. Hence, the generalization of our ECG-GLS score prediction to all strain vendors requires further investigation. Finally, the validation cohort included patients with HF, which made the predictive ability of our ECG-GLS score lower than expected. Further studies are needed to demonstrate the utility of the ECG-GLS score in various populations.

### Conclusions

The ECG-GLS score, estimated from ECG features using an AI-based algorithm, shows a strong correlation with the LVGLS measured on echocardiography and is effective in risk stratification for the long-term prognosis after acute HF. These findings suggest the potential role of the AI-based ECG algorithm as a practical alternative to the LVGLS on echocardiography.

## CLINICAL PERSPECTIVES

**Competency in Patient Care and Procedural Skills:** The ECG-GLS score can be used to predict the value of the LVGLS, diagnose LV systolic dysfunction, and forecast the outcome of patients with heart failure more easily, reducing time and costs.

**Translational Outlook:** AI can be used to expand the capabilities of ECG to rival state-of-the-art technology in other modalities, such as the LVGLS on echocardiography.

## Data Availability

All data produced in the present study are available upon reasonable request to the authors

## Acknowledgements

None

## ABBREVIATIONS

ACD: all-cause death
AI: artificial intelligence
AUROC: area under the receiver-operating characteristic curve
ECG: electrocardiography
EF: ejection fraction
GLS: global longitudinal strain
HCM: hypertrophic cardiomyopathy
HF: heart failure
HFrEF: heart failure with reduced ejection fraction
HHF: hospitalization for heart failure
LV: left ventricle
LVEF: left ventricular ejection fraction
LVGLS: left ventricular global longitudinal strain

## References

1. Potter E, Marwick TH. Assessment of Left Ventricular Function by Echocardiography: The Case for Routinely Adding Global Longitudinal Strain to Ejection Fraction. JACC Cardiovasc Imaging 2018;11:260–274.

2. Park JJ, Park JB, Park JH, Cho GY. Global Longitudinal Strain to Predict Mortality in Patients With Acute Heart Failure. J Am Coll Cardiol 2018;71:1947–1957.

3. Magne J, Cosyns B, Popescu BA et al. Distribution and Prognostic Significance of Left Ventricular Global Longitudinal Strain in Asymptomatic Significant Aortic Stenosis: An Individual Participant Data Meta-Analysis. JACC Cardiovasc Imaging 2019;12:84–92.

4. Zhu D, Ito S, Miranda WR et al. Left Ventricular Global Longitudinal Strain Is Associated With Long-Term Outcomes in Moderate Aortic Stenosis. Circ Cardiovasc Imaging 2020;13:e009958.

5. Alashi A, Mentias A, Abdallah A et al. Incremental Prognostic Utility of Left Ventricular Global Longitudinal Strain in Asymptomatic Patients With Significant Chronic Aortic Regurgitation and Preserved Left Ventricular Ejection Fraction. JACC Cardiovasc Imaging 2018;11:673–682.

6. Tower-Rader A, Mohananey D, To A, Lever HM, Popovic ZB, Desai MY. Prognostic Value of Global Longitudinal Strain in Hypertrophic Cardiomyopathy: A Systematic Review of Existing Literature. JACC Cardiovasc Imaging 2019;12:1930–1942.

7. Siontis KC, Noseworthy PA, Attia ZI, Friedman PA. Artificial intelligence-enhanced electrocardiography in cardiovascular disease management. Nat Rev Cardiol 2021;18:465–478.

8. Attia ZI, Harmon DM, Behr ER, Friedman PA. Application of artificial intelligence to the electrocardiogram. Eur Heart J 2021;42:4717–4730.

9. Choi YJ, Park MJ, Ko Y et al. Artificial intelligence versus physicians on interpretation of printed ECG images: Diagnostic performance of ST-elevation myocardial infarction on electrocardiography. Int J Cardiol 2022;363:6–10.

10. Adedinsewo D, Carter RE, Attia Z et al. Artificial Intelligence-Enabled ECG Algorithm to Identify Patients With Left Ventricular Systolic Dysfunction Presenting to the Emergency Department With Dyspnea. Circ Arrhythm Electrophysiol 2020;13:e008437.

11. Moon MG, Hwang IC, Lee HJ et al. Reverse Remodeling Assessed by Left Atrial and Ventricular Strain Reflects Treatment Response to Sacubitril/Valsartan. JACC Cardiovasc Imaging 2022;15:1525–1541.

12. Kim D, Hwang JE, Cho Y et al. A Retrospective Clinical Evaluation of an Artificial Intelligence Screening Method for Early Detection of STEMI in the Emergency Department. J Korean Med Sci 2022;37:e81.

13. Lang RM, Badano LP, Mor-Avi V et al. Recommendations for cardiac chamber quantification by echocardiography in adults: an update from the American Society of Echocardiography and the European Association of Cardiovascular Imaging. J Am Soc Echocardiogr 2015;28:1–39 e14.

14. Attia ZI, Noseworthy PA, Lopez-Jimenez F et al. An artificial intelligence-enabled ECG algorithm for the identification of patients with atrial fibrillation during sinus rhythm: a retrospective analysis of outcome prediction. The Lancet 2019;394:861–867.

15. Attia ZI, Kapa S, Lopez-Jimenez F et al. Screening for cardiac contractile dysfunction using an artificial intelligence-enabled electrocardiogram. Nat Med 2019;25:70–74.

16. Kwon JM, Kim KH, Eisen HJ et al. Artificial intelligence assessment for early detection of heart failure with preserved ejection fraction based on electrocardiographic features. Eur Heart J Digit Health 2021;2:106–116.

17. Ko WY, Siontis KC, Attia ZI et al. Detection of Hypertrophic Cardiomyopathy Using a Convolutional Neural Network-Enabled Electrocardiogram. J Am Coll Cardiol 2020;75:722–733.

18. Tison GH, Zhang J, Delling FN, Deo RC. Automated and Interpretable Patient ECG Profiles for Disease Detection, Tracking, and Discovery. Circulation: Cardiovascular Quality and Outcomes 2019;12.

19. Tison GH, Siontis KC, Abreau S et al. Assessment of Disease Status and Treatment Response With Artificial Intelligence-Enhanced Electrocardiography in Obstructive Hypertrophic Cardiomyopathy. J Am Coll Cardiol 2022;79:1032–1034.

20. Lee E, Ito S, Miranda WR et al. Artificial intelligence-enabled ECG for left ventricular diastolic function and filling pressure. NPJ Digit Med 2024;7:4.

21. Farsalinos KE, Daraban AM, Unlu S, Thomas JD, Badano LP, Voigt JU. Head-to-Head Comparison of Global Longitudinal Strain Measurements among Nine Different Vendors: The EACVI/ASE Inter-Vendor Comparison Study. J Am Soc Echocardiogr 2015;28:1171–1181, e2.

22. Voigt JU, Pedrizzetti G, Lysyansky P et al. Definitions for a common standard for 2D speckle tracking echocardiography: consensus document of the EACVI/ASE/Industry Task Force to standardize deformation imaging. J Am Soc Echocardiogr 2015;28:183–93.

