## Supplementary file for "ECG-derived global longitudinal strain using artificial intelligence: A comparative study with transthoracic echocardiography"

Supplementary Figure 1. Precision-recall curve for predicting LVGLS ≤12% using the ECG-GLS score.

Supplementary table 1. Univariable Cox Proportional Hazard Regression Analysis for 5-Year All-Cause Death

Supplementary Table 2. Univariable Cox Proportional Hazard Regression Analysis for 5-Year All-Cause Death and Hospitalization for Heart Failure

**Supplementary Figure 1.** Precision-recall curve for predicting LVGLS ≤12% using the ECG-GLS score.

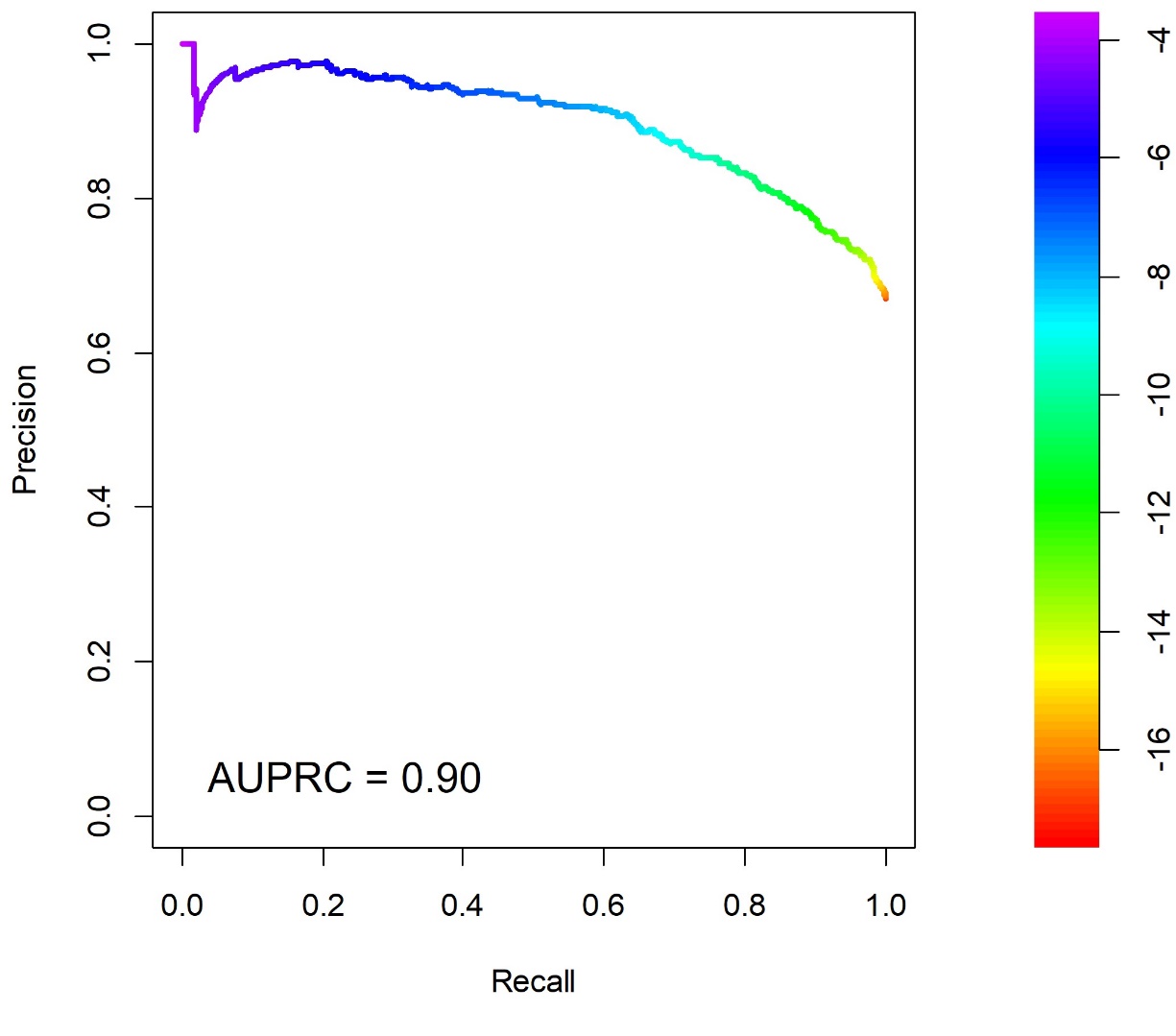

Abbreviations: AUPRC, area under the precision-recall curve; ECG-GLS, electrocardiography-derived global longitudinal strain; LVGLS, left ventricular global longitudinal strain on echocardiography

**Supplementary Table 1. Univariable Cox Proportional Hazard Regression Analysis for 5-Year All-Cause Death**

|  | HR | 95% CI | P value |
| --- | --- | --- | --- |
| Age (years) | 1.03 | 1.03-1.04 | <0.001 |
| Age >75 years | 2.01 | 1.69-2.39 | <0.001 |
| Male sex | 1.04 | 0.88-1.24 | 0.623 |
| SBP (mmHg) | 1.00 | 0.99-1.00 | 0.011 |
| DBP (mmHg) | 0.99 | 0.98-0.99 | <0.001 |
| Heart rate (bpm) | 1.00 | 1.00-1.00 | 0.054 |
| DM | 1.18 | 0.99-1.42 | 0.072 |
| Hypertension | 0.94 | 0.79-1.11 | 0.455 |
| Prior CAD | 0.83 | 0.69-1.01 | 0.057 |
| Atrial fibrillation | 0.87 | 0.73-1.05 | 0.142 |
| BUN (mg/dL) | 1.01 | 1.01-1.02 | <0.001 |
| Creatinine (mg/dL) | 1.04 | 1.01-1.08 | 0.026 |
| B-type natriuretic peptide (pg/mL) | 1.00 | 1.00-1.00 | <0.001 |
| Beta-blocker | 0.62 | 0.52-0.76 | <0.001 |
| RAS blocker | 0.68 | 0.57-0.81 | <0.001 |
| Mineralocorticoid receptor antagonist | 0.96 | 0.79-1.16 | 0.675 |
| LV end-diastolic dimension (mm) | 0.99 | 0.98-1.00 | 0.007 |
| LV end-systolic dimension (mm) | 0.99 | 0.98-1.00 | 0.079 |
| Right ventricular systolic pressure (mmHg) | 1.01 | 1.00-1.02 | 0.003 |
| E/e' | 1.02 | 1.04-1.03 | <0.001 |
| E/e' >15 | 1.50 | 1.22-1.85 | <0.001 |
| LVEF (%) | 1.10 | 0.99-1.00 | 0.130 |
| LVEF ≤30% | 1.18 | 0.98-1.41 | 0.083 |
| LVGLS (%) | 0.96 | 0.94-0.97 | <0.001 |
| LVGLS ≤12% | 1.41 | 1.16-1.71 | <0.001 |
| ECG-GLS score | 0.94 | 0.92-0.97 | <0.001 |
| ECG-GLS score ≤12 | 1.36 | 1.12-1.66 | 0.002 |

Abbreviations as in Table 1 and 2.

**Supplementary Table 2. Univariable Cox Proportional Hazard Regression Analysis for 5-Year All-Cause Death and Hospitalization for Heart Failure**

|  | HR | 95% CI | P value |
| --- | --- | --- | --- |
| Age (years) | 1.03 | 1.02-1.03 | <0.001 |
| Age >75 years | 1.92 | 1.67-2.21 | <0.001 |
| Male sex | 1.03 | 0.90-1.19 | 0.636 |
| SBP (mmHg) | 1.00 | 1.00-1.00 | 0.215 |
| DBP (mmHg) | 0.99 | 0.99-1.00 | <0.001 |
| Heart rate (bpm) | 1.00 | 1.00-1.01 | 0.017 |
| Diabetes mellitus | 1.35 | 1.17-1.56 | <0.001 |
| Hypertension | 0.82 | 0.71-0.94 | 0.004 |
| Ischemic heart disease | 0.77 | 0.66-0.90 | <0.001 |
| Atrial fibrillation | 0.90 | 0.77-1.04 | 0.138 |
| BUN (mg/dL) | 1.01 | 1.01-1.02 | <0.001 |
| Creatinine (mg/dL) | 1.04 | 1.01-1.07 | 0.008 |
| B-type natriuretic peptide (pg/mL) | 1.00 | 1.00-1.00 | 0.002 |
| Beta-blocker | 0.80 | 0.69-0.93 | 0.003 |
| RAS blocker | 0.83 | 0.72-0.95 | 0.009 |
| Mineralocorticoid receptor antagonist | 0.96 | 0.82-1.12 | 0.596 |
| LV end-diastolic dimension (mm) | 0.99 | 0.98-1.00 | 0.019 |
| LV end-systolic dimension (mm) | 0.99 | 0.99-1.00 | 0.031 |
| Right ventricular systolic pressure (mmHg) | 1.01 | 1.00-1.02 | 0.011 |
| E/e' | 1.02 | 1.01-1.02 | <0.001 |
| E/e' >15 | 1.43 | 1.21-1.68 | <0.001 |
| LVEF (%) | 1.00 | 0.99-1.00 | 0.430 |
| LVEF ≤30% | 1.07 | 0.92-1.25 | 0.351 |
| LVGLS (%) | 0.97 | 0.96-0.98 | <0.001 |
| LVGLS ≤12% | 1.28 | 1.10-1.48 | 0.002 |
| ECG-GLS score | 0.96 | 0.93-0.98 | <0.001 |
| ECG-GLS score ≤12 | 1.31 | 1.12-1.53 | <0.001 |

Abbreviations as in Table 1 and 2.
